# Effectiveness of physical therapy for locomotive syndrome: Study protocol for a systematic review and meta-analysis

**DOI:** 10.1101/2024.11.08.24317013

**Authors:** Chadapa Rungruangbaiyok, Hiroyuki Ohtsuka, Charupa Lektip, Jiraphat Nawarat, Eiji Miyake, Keiichiro Aoki, Yasuko Inaba, Yoshinori Kagaya

## Abstract

Locomotive syndrome, characterized by impaired mobility due to musculoskeletal disorders, poses a significant public health challenge, especially in the aging population. Locomotive syndrome limits physical activity, increases fall risk, leads to dependency, and diminishes quality of life. Effective interventions are urgently required. This systematic review and meta-analysis aimed to evaluate the effectiveness of physical therapy in improving the symptoms of locomotive syndrome. A systematic evaluation of its effectiveness compared with other interventions is crucial for informing clinical practice and policy decisions. This systematic review and meta-analysis follows the PRISMA-P guidelines. Studies involving individuals diagnosed with locomotive syndrome, without restrictions on age, sex, or location, were included. The interventions that are reviewed encompass exercise programs, manual therapy, neuromuscular stimulation, and balance training, which are be compared with no intervention or alternative therapies. The primary outcomes include improvements in functional mobility and physical performance, a reduction in symptoms, and the progression of locomotive syndrome. Secondary outcomes include adherence to therapy, safety, quality of life, patient satisfaction, and psychological well-being. Randomized controlled and non-randomized controlled trials published in English were searched from PubMed, CENTRAL, CINAHL, PEDro, Ichushi Web, and the Thai-Journal Citation Index Center. Independent reviewers performed data extraction and assess the risk of bias. A meta-analyses was conducted using RevMan 5.4 software, with subgroup analyses to address heterogeneity. The Grading of Recommendations Assessment, Development and Evaluation (GRADE) approach was used to evaluate the certainty of evidence. This review aims to provide robust evidence on the effectiveness of physical therapy in managing locomotive syndrome and to guide clinical practice and healthcare policy decisions.

## Introduction

Locomotive syndrome (LS), a condition characterized by impaired mobility due to musculoskeletal disorders, has become a significant public health concern, particularly in the aging population [1–4]. This syndrome, which encompasses a range of conditions, such as osteoarthritis, sarcopenia, and osteoporosis, adversely affects the quality of life by limiting physical activity and increasing the risk of falls and dependency [1,2,5,6].

The increasing prevalence of LS, particularly in aging populations, underscores the urgent need for effective interventions to mitigate its impact [1,2,5,7,8]. In Japan, where the concept of LS was first introduced, the aging population was particularly affected. The prevalence of LS among older adults is rising, with a corresponding increase in the risk of falls and fractures, which further exacerbates the associated disability and healthcare costs [1,2,4,5,7–10]. Other countries with aging populations are experiencing similar challenges; this trend is also not unique to Japan [11–13].

Given these trends, there is an urgent need for effective interventions to prevent or slow the progression of LS. Physical therapy interventions, through a combination of strength training, balance exercises, gait training, pain management, and flexibility exercises, provide a comprehensive approach to managing LS. The robust evidence from several studies underscores the effectiveness of physical therapy in preventing or slowing the progression of this condition, thereby enhancing the quality of life and reducing healthcare costs associated with disability and dependency [10,14–24]. However, to ensure the optimal allocation of healthcare resources and the best outcomes for patients, it is critical to systematically evaluate the effectiveness of physical therapy compared with other available interventions.

Although physical therapy is commonly prescribed to address the symptoms of LS [2–4,18,19], its relative effectiveness compared to other treatments, such as pharmacological approaches, surgical options, or alternative therapies, remains a subject of ongoing debate [9,18,20,27–30]. Although LS encompasses distinct conditions such as osteoarthritis, osteoporosis, and sarcopenia, which affect various organ systems, these conditions share common pathophysiological mechanisms that lead to impaired mobility and increased fall risk. Therefore, grouping these conditions together allows for a comprehensive evaluation of interventions aimed at improving overall locomotor function. This systematic review and meta-analysis aim to evaluate the effectiveness of physical therapy interventions in improving functional mobility and overall health-related quality of life in individuals with LS. We hypothesize that physical therapy is more effective than other treatment modalities (e.g., pharmacological, surgical, or alternative therapies) in improving functional mobility and health-related quality of life in individuals with LS. The clinical relevance of physical therapy interventions will be assessed by measuring changes in functional mobility and health-related quality of life that meet or exceed established minimal clinically important differences (MCIDs), indicating a meaningful impact on patient function and daily activities. To account for potential biases related to age, sex, and frailty, subgroup analyses will be performed. These analyses will help determine whether different demographic or clinical characteristics influence the effectiveness of physical therapy in managing LS.

## Methods

This systematic review was conducted and reported in accordance with the Preferred Reporting Items for Systematic Reviews and Meta-Analyses Protocols (PRISMA-P) statement [30]. This protocol has been registered with the International Prospective Register of Systematic Reviews (PROSPERO) database under the registration number (CRD42024515983). Adherence to the PRISMA-P guidelines ensures that our review process is methodologically rigorous and transparent, thus providing a reliable foundation for synthesizing existing research (Supplemental Appendix 1). This study has not yet started. Data collection will begin following the acceptance of this protocol, using the developed search strategy. The data collection process is expected to be completed within six months from the start.

### Participants/population

This systematic review includes individuals diagnosed with LS [1–4], without restrictions on age, sex, or geographical location. Individuals not diagnosed with LS or those with other diagnoses were excluded from the review. This approach ensures a focused and comprehensive analysis of the population affected by LS.

### Interventions/exposures

This systematic review examines the various physical therapy interventions in patients with LS. Interventions include exercise programs, manual therapy, neuromuscular stimulation, and balance training. Each of these interventions was analyzed for their effectiveness in managing and improving the symptoms and overall function of individuals diagnosed with LS.

### Comparators/controls

The comparators do not include interventions or other interventions (active controls). This approach allows us to assess the relative effectiveness of various physical therapy interventions for LS by comparing them with both a lack of intervention and alternative therapeutic strategies.

### Main outcomes

The primary outcomes of this systematic review includes the following:

- Improvement in Functional Mobility: This was assessed through measures of enhanced mobility, such as walking speed, gait analysis, and the ability to perform daily activities.
- Physical Performance Measures: Objective assessments, such as balance, strength, and endurance tests, were used to evaluate physical performance.
- Reduction in Symptoms: This includes a decrease in pain levels, stiffness, and other symptoms commonly associated with LS.
- Progression of LS: Any changes in the progression or severity of LS over time were also tracked and analyzed.

### Additional outcomes

The secondary outcomes of this systematic review includes the following:

- Adherence to Therapy: The extent to which patients adhere to their prescribed physical therapy regimens was evaluated.
- Safety and Adverse Events: Documentation and analysis of any adverse events or safety concerns associated with the physical therapy interventions was included.
- Quality of Life: Assessment of overall well-being and quality of life improvements as perceived by the patients was conducted.
- Patient Satisfaction: Patient-reported satisfaction with the physical therapy treatment and its outcomes were considered.
- Psychological Well-being: The impact of physical therapy on the psychological and emotional well-being of patients was examined.

### Eligibility criteria

This systematic review includes randomized controlled trials (RCTs) and non-RCTs published in English with full-text availability. Review articles, conference abstracts, and letters to the editor were excluded. These criteria ensured that the review focuses on high-quality peer-reviewed studies that provide comprehensive data for rigorous analysis and synthesis while excluding sources that typically lack detailed data.

### Information sources and search strategy

The following databases will be used for the search: PubMed, Cochrane Central Register of Controlled Trials (CENTRAL), CINAHL, PEDro, Scopus, Ichushi Web (in Japanese), and Thai Journal Citation Index Center (in Thailand). The search will be limited to studies published in English, Japanese, and Thai focusing on human subjects. Abstracts and conference proceedings will be also included, and the authors will be contacted for additional details, if necessary. This comprehensive search strategy ensures a thorough and inclusive collection of relevant studies for the systematic review. The primary search terms will be defined as: ‘locomotive syndrome,’ ‘rehabilitation,’ ‘physical therapy,’ ‘exercise,’ ‘electrical stimulation,’ and ‘clinical trial.’ A preliminary version of the search strategy for the relevant databases will be included in the supplementary material (Supplemental Appendix 2).

### Data extraction (selection and coding)

This review will be conducted following the PRISMA guidelines. Searches will be performed by two independent reviewers (HO and CR) using widely recognized databases. The search terms included a combination of MeSH (Medical Subject Headings) terms from these databases and free-text search terms agreed upon by all authors. Rayyan was used to manage the studies across different databases.

Two independent reviewers (HO, CR, CL, JN, EM, YI, and YK) will assess the titles and abstracts of the studies to determine their eligibility based on the inclusion criteria. Studies that could not be conclusively evaluated based on the title and abstract alone were further appraised by reviewing the full text. In cases of disagreement between the two reviewers, a third reviewer will be available for discussion to resolve the issue.

For data extraction, two independent reviewers (HO and CR) gathered detailed information on the study design and methodology, demographic and baseline characteristics of the participants, sample size, and measures of effect. Any discrepancies in judgment between the reviewers was resolved through discussion with a third reviewer. Any missing data in the articles will be requested from the study authors as necessary. The data extraction process will be meticulously documented and organized using a standard Microsoft Excel spreadsheet.

### Risk of bias assessment

The Risk of bias in the RCTs will be assessed using the Cochrane Risk of Bias tool (RoB 2.0). Two independent reviewers (HO and CR) will critically assess all the included studies. The evaluation includes the following items.

- Bias arising from the randomization process.
- Bias due to deviations from the intended intervention.
- Bias due to missing outcome data.
- Bias in the measurement of outcome.
- Bias in the selection of the reported result.
- Overall effect.

For each item, each study will be evaluated as having a low, uncertain, or high risk of bias. Any discrepancies between the reviewers were discussed and resolved by a third reviewer, if necessary.

The risk of bias in non-RCTs will be assessed using the Risk of Bias Assessment Tool for Nonrandomized Studies (RoBANS). Two independent reviewers (HO and CR) will critically assess all the included studies. The evaluation includes the following items.

- Selection of participants.
- Confounding variables.
- Measurement of exposure.
- Blinding of the outcome assessment.
- Incomplete outcome data.
- Selective outcome reporting.

For each item, each study will be evaluated as having a low, unclear, or high risk of bias. Any discrepancies between the reviewers were be discussed and resolved by a third reviewer, if necessary.

### Data synthesis

If numerous RCTs consistently corroborate their findings, a meta-analysis will be conducted. For this process, RevMan 5.4 software will be employed. For the analysis of continuous data, weighted mean differences (MD), including means and standard deviations, will be utilized. The standardized mean difference (SMD) will be applied to coalesce multiple measurements of identical outcome variables. To derive aggregate estimates, a random-effects model will be adopted, accompanied by forest plots to graphically represent the findings. The I² test will be used to assess heterogeneity; a value surpassing 50% in the I² test indicates substantial heterogeneity, necessitating the execution of a subgroup analysis. Consequently, a comprehensive table summarizing the results will be compiled in alignment with the reported findings.

### Subgroup analysis

When multiple trials are present within each subgroup, the analyses will be stratified based on participant demographics, including sex (male/female), nature of the intervention, and type of control group (either no intervention or active control). Furthermore, subgroup analyses will be conducted in cases with a significant degree of heterogeneity. These analyses may help identify variations in treatment effects across different participant characteristics and intervention types, providing a more nuanced understanding of the data.

### Assessment of certainty of evidence

The certainty of evidence will be evaluated using the Grading of Recommendations, Assessment, Development, and Evaluations (GRADE) approach. This systematic method will be used to assess the quality of evidence across the domains of risk of bias, consistency of effects, imprecision, indirectness, and publication bias.

Each outcome will be rated as high, moderate, low, or very low. The initial rating for RCTs will be high; however, it might be downgraded based on the aforementioned domains. Conversely, the rating for observational studies started as low but was upgraded if the evidence showed a large effect, a dose-response gradient, or if all plausible biases reduced an apparent treatment effect.

Two independent reviewers (HO and CR) will be conducted the GRADE assessment, and any discrepancies will be resolved through discussion or consultation with a third reviewer. The results of the GRADE assessment will be summarized in a table that provided a clear and transparent evaluation of the certainty of the evidence for each outcome.

### Patient and public involvement

There will be no patient or public involvement in the design, conduct, reporting, or dissemination of this systematic review.

### Ethical considerations

No ethical approval will be required for this systematic review, as it involved the analysis of data from previously published studies and did not involve direct contact with patients or the collection of primary data.

## Discussion

Several studies have explored the benefits of physical therapy for managing musculoskeletal conditions, thereby providing a foundation for our research. For instance, the meta-analysis by Liu and Latham (2009) highlighted the effectiveness of progressive resistance strength training in improving physical function in older adults [17]. Sherrington et al. (2011) conducted a meta-analysis demonstrating the benefits of exercise in preventing falls among older adults [18]. Furthermore, Fransen et al. (2015) reviewed the impact of exercise on knee osteoarthritis and found significant improvements in pain and physical function [19]. Despite these findings, the relative effectiveness of various physical therapy interventions, specifically for LS, remains unclear. Our research aimed to fill this gap by systematically evaluating and synthesizing existing evidence and providing a comprehensive assessment of the impact of physical therapy on this condition.

Our systematic review and meta-analysis has several strengths that enhanced the reliability and applicability of our findings. First, we used a rigorous and transparent methodology adhering to the PRISMA-P guidelines, which ensured a systematic and unbiased approach to data collection, extraction, and analysis. The inclusion of both RCTs and non-RCTs allowed for a comprehensive evaluation of the evidence. Second, our extensive search strategy that included multiple databases and languages (English, Japanese, and Thai), ensured a thorough and inclusive collection of relevant studies, thereby minimizing the risk of publication bias. Third, we employed robust statistical methods, such as random-effects models and subgroup analyses, to account for heterogeneity and provide a nuanced understanding of treatment effects across different populations and intervention types. Additionally, the use of Cochrane’s risk-of-bias tool and the GRADE approach to assessed the certainty of evidence that further strengthened the credibility of our findings.

Several practical and operational challenges were anticipated in conducting this study. Dealing with the heterogeneity of the included studies, such as variations in study design, intervention types, and outcome measures, should be carefully considered. This was addressed through subgroup analyses and the use of random-effect models to account for variability. Efficient data extraction and management are critical when employing standardized forms and software, such as Rayyan, for study screening. Assessing the risk of bias and ensuring accurate data extraction required multiple reviewers and a rigorous validation process.

When interpreting the results, it is important to distinguish between statistically significant differences and clinically meaningful outcomes. Even if a result is statistically significant, it does not necessarily translate into a clinically relevant improvement. Therefore, we used minimal clinically important differences (MCIDs) as a benchmark to determine whether the observed changes in functional mobility and quality of life were clinically significant. This ensures that the results not only demonstrate statistical validity but also have practical implications for patient care. For example, a small, statistically significant improvement in gait speed might not meet the threshold for MCID, meaning that while the intervention had an effect, it may not be sufficient to make a meaningful difference in a patient’s daily life. This approach allows clinicians to base treatment decisions on changes that are both statistically supported and clinically relevant, ensuring that the interventions offer real-world benefits to patients.

Adherence to physical therapy is another significant factor that can influence patient outcomes. We evaluated the adherence rates and explored strategies to improve adherence, considering their feasibility and impact on study outcomes. In addition, understanding the long-term sustainability of physical therapies is crucial. We included follow-up studies to assess the duration of the observed effects and provide a comprehensive understanding of the long-term effectiveness of physical therapy.

## Conclusion

This systematic review and meta-analysis aimed to provide compelling evidence supporting the effectiveness of physical therapy for managing LS. By addressing the practical and operational challenges and employing a rigorous and comprehensive methodology, our research contributed to the optimization of physical therapy interventions for this condition. These findings may guide clinical practice, healthcare providers in making evidence-based decisions, and ultimately enhance the quality of life of individuals with LS.

## Data Availability

Deidentified research data will be made publicly available when the study is completed and published.

## Acknowledgments

We thank Ms. Tomoko Morimasa and Ms. Asae Ito, (librarian, Showa University) for their advice on creating the search strategy. Finally, we would like to thank Editage (www.editage.com) for the English language editing.

## Author Contributions

**Conceptualization**: Chadapa Rungruangbaiyok, Hiroyuki Ohtsuka, and Yoshinori Kagaya

**Data curation**: Chadapa Rungruangbaiyok and Hiroyuki Ohtsuka

**Formal analysis**: Chadapa Rungruangbaiyok and Hiroyuki Ohtsuka

**Funding Acquisition:** Chadapa Rungruangbaiyok, Hiroyuki Ohtsuka, Eiji Miyake, Keiichiro Aoki, Yasuko Inaba, and Yoshinori Kagaya

**Investigation**: Chadapa Rungruangbaiyok and Hiroyuki Ohtsuka

**Methodology**: Hiroyuki Ohtsuka

**Project administration**: Chadapa Rungruangbaiyok, Hiroyuki Ohtsuka, and Yoshinori Kagaya

**Resources:** Hiroyuki Ohtsuka

**Software:** Chadapa Rungruangbaiyok and Hiroyuki Ohtsuka

**Supervision:** Chadapa Rungruangbaiyok, Hiroyuki Ohtsuka, Charupa Lektip, Jiraphat Nawarat, Eiji Miyake, Keiichiro Aoki, Yasuko Inaba, and Yoshinori Kagaya

**Validation:** Chadapa Rungruangbaiyok, Hiroyuki Ohtsuka, Charupa Lektip, Jiraphat Nawarat, Eiji Miyake, Keiichiro Aoki, Yasuko Inaba, and Yoshinori Kagaya.

**Visualization:** Chadapa Rungruangbaiyok, Hiroyuki Ohtsuka, Charupa Lektip, Jiraphat Nawarat, Eiji Miyake, Keiichiro Aoki, Yasuko Inaba, and Yoshinori Kagaya

**Writing – original draft:** Chadapa Rungruangbaiyok and Hiroyuki Ohtsuka

**Writing – review & editing:** Chadapa Rungruangbaiyok, Hiroyuki Ohtsuka, Charupa Lektip, Jiraphat Nawarat, Eiji Miyake, Keiichiro Aoki, Yasuko Inaba, and Yoshinori Kagaya

## References

1. Nakamura K. A “super-aged” society and the “locomotive syndrome”. J Orthop Sci. 2008;13: 1–2. doi: 10.1007/s00776-007-1202-6.

2. Seichi A, Hoshino Y, Doi T, Akai M, Tobimatsu Y, Iwaya T. Development of a screening tool for risk of locomotive syndrome in the elderly: The 25-question Geriatric Locomotive Function Scale. J Orthop Sci. 2012;17: 163–172. doi: 10.1007/s00776-011-0193-5.

3. Yoshimura N, Muraki S, Oka H, Tanaka S, Ogata T, Kawaguchi H, Akune T, Nakamura K. Association between new indices in the locomotive syndrome risk test and decline in mobility: Third survey of the ROAD study. J Orthop Sci. 2015;20:896–905. doi: 10.1007/s00776-015-0741-5.

4. Kataoka H, Miyatake N, Ichikawa H, Arakawa Y, Mori Y. Relationship of locomotive syndrome with health-related quality of life among patients with obstructive sleep apnea syndrome. J Phys Ther Sci. 2017;29:1129–1133. doi: 10.1589/jpts.29.1129.

5. Muraki S, Akune T, Oka H, En-Yo Y, Yoshida M, Saika A, Suzuki T, Yoshida H, Ishibashi H, Tokimura F, Yamamoto S, Nakamura K, Kawaguchi H, Yoshimura N. Impact of knee and low back pain on health-related quality of life in Japanese women: The Research on Osteoarthritis Against Disability (ROAD). Mod Rheumatol. 2010;20:444–451. doi: 10.1007/s10165-010-0307-5.

6. Cruz-Jentoft AJ, Baeyens JP, Bauer JM, Boirie Y, Cederholm T, Landi F, et al. Sarcopenia: European consensus on definition and diagnosis. Age Ageing. 2010;39: 412–423. doi: 10.1093/ageing/afq034.

7. Ishibashi H. Locomotive syndrome in Japan. Osteoporos Sarcopenia. 2018;4(3):86–94. doi: 10.1016/j.afos.2018.09.004.

8. Taniguchi M, Ikezoe T, Tsuboyama T, Tabara Y, Matsuda F, Ichihashi N. Prevalence and physical characteristics of locomotive syndrome stages as classified by the new 2020 criteria in older Japanese people: Results from the Nagahama study. BMC Geriatr. 2021;21:1–9. doi: 10.1186/s12877-021-02440-2.

9. Muraki S, Akune T, Oka H, En-Yo Y, Yoshida M, Nakamura K, Kawaguchi H, Yoshimura N. Prevalence of falls and association with knee osteoarthritis, lumbar spondylosis, and knee and lower back pain in Japanese men and women. Arthritis Care Res (Hoboken). 2011;63(10):1425–1431. doi: 10.1002/acr.20562.

10. Nakamura K. The concept and treatment of locomotive syndrome: Its acceptance and spread in Japan. J Orthop Sci. 2011;16: 489–491. doi: 10.1007/s00776-011-0108-5.

11. Cieza A, Causey K, Kamenov K, Hanson SW, Chatterji S, Vos T. Global estimates of the need for rehabilitation based on the Global Burden of Disease Study 2019: A systematic analysis for the Global Burden of Disease Study 2019. Lancet. 2021;396(10267):2006–2017. doi: 10.1016/S0140-6736(20)32340-0.

12. Beard JR, Officer A, de Carvalho IA, Sadana R, Pot AM, Michel JP, Peeters GM. The World report on ageing and health: A policy framework for healthy ageing. Lancet. 2016;387(10033):2145–2154. doi: 10.1016/S0140-6736(15)00516-4.

13. Dennison EM, Mohamed MA, Cooper C. Epidemiology of osteoporosis. Rheum Dis Clin North Am. 2006;32(4):617–629. doi: 10.1016/j.rdc.2006.08.003.

14. Peeters G, van Schoor NM, Lips P. Fall risk: The clinical relevance of falls and how to integrate fall risk with fracture risk. Best Pract Res Clin Rheumatol. 2009;23: 797–804. doi: 10.1016/j.berh.2009.09.004.

15. Clynes M, Edwards M, Buehring B, Dennison E, Binkley N, Cooper C. Definitions of sarcopenia: Associations with previous falls and fracture in a population sample. Calcif Tissue Int. 2015;97(5):445–452. doi: 10.1007/s00223-015-0044-z.

16. Prince MJ, Wu F, Guo Y, Gutierrez Robledo LM, O’Donnell M, Sullivan R, et al. The burden of disease in older people and implications for health policy and practice. Lancet. 2015;385: 549–562. doi: 10.1016/S0140-6736(14)61347-7.

17. Liu CJ, Latham NK. Progressive resistance strength training for improving physical function in older adults. Cochrane Database Syst Rev. 2009;3. doi: 10.1002/14651858.CD002759.pub2.

18. Sherrington C, Tiedemann A, Fairhall N, Close JC, Lord SR. Exercise to prevent falls in older adults: An updated meta-analysis and best practice recommendations. N S W Public Health Bull. 2011;22: 78–83. doi: 10.1071/NB10056.

19. Fransen M, McConnell S, Harmer AR, Van der Esch M, Simic M, Bennell KL. Exercise for osteoarthritis of the knee: A Cochrane systematic review. Br J Sports Med. 2015;49: 1554–1557. doi: 10.1136/bjsports-2015-095424.

20. Brosseau L, Wells GA, Pugh AG, Smith CA, Rahman P, Suarez-Almazor ME. Ottawa Panel evidence-based clinical practice guidelines for therapeutic exercises and manual therapy in the management of osteoarthritis. Phys Ther. 2014;94: 1017–1025. doi: 10.2522/ptj.20130329.

21. American College of Sports Medicine (ACSM). Exercise and physical activity for older adults. Med Sci Sports Exerc. 2009;41(7):1510–1530. doi: 10.1249/MSS.0b013e3181a0c95c.

22. Bennell KL, Dobson F, Hinman RS. Exercise in osteoarthritis: Moving from prescription to adherence. Best Pract Res Clin Rheumatol. 2014;28: 93–117. doi: 10.1016/j.berh.2014.01.009.

23. Pisters MF, Veenhof C, van Meeteren NL, Ostelo RW, de Bakker DH, Dekker J. Long-term effectiveness of exercise therapy in patients with osteoarthritis of the hip or knee: A systematic review. Arthritis Care Res (Hoboken). 2010;62(7):1087–1096. doi: 10.1002/acr.20182.

24. Akune T, Muraki S, Oka H, Tanaka S, Kawaguchi H, Nakamura K, Yoshimura N. Exercise habits during middle age are associated with lower prevalence of sarcopenia: The ROAD study. Osteoporos Int. 2014;25(3):1081–1088. doi: 10.1007/s00198-013-2550-z.

25. Muraki S, Akune T, Oka H, Mabuchi A, En-Yo Y, Yoshida M, Yoshimura N. Association of occupational activity with radiographic knee osteoarthritis and lumbar spondylosis in elderly patients of population-based cohorts: A large-scale population-based study. Arthritis Rheum. 2011;63(2):400–407. doi: 10.1002/art.30108.

26. Skou ST, Roos EM, Laursen MB, Rathleff MS, Arendt-Nielsen L, Simonsen O, Rasmussen S. A randomized, controlled trial of total knee replacement. N Engl J Med. 2015;373(17):1597–1606. doi: 10.1056/NEJMoa1505467.

27. Foster NE, Anema JR, Cherkin D, Chou R, Cohen SP, Gross DP, Maher CG. Prevention and treatment of low back pain: Evidence, challenges, and promising directions. Lancet. 2018;391(10137):2368–2383. doi: 10.1016/S0140-6736(18)30489-6.

28. Mafi JN, McCarthy EP, Davis RB, Landon BE. Worsening trends in the management and treatment of back pain. JAMA Intern Med. 2013;173: 1573–1581. doi: 10.1001/jamainternmed.2013.8992.

29. Artus M, Jordan KP, Croft PR. The clinical course of low back pain: A meta-analysis comparing outcomes in randomized clinical trials (RCTs) and observational studies. BMC Musculoskelet Disord. 2014;15:68. doi: 10.1186/1471-2474-15-68.

30. Page MJ, McKenzie JE, Bossuyt PM, Boutron I, Hoffmann TC, Mulrow CD, et al. The PRISMA 2020 statement: An updated guideline for reporting systematic reviews. PLOS Med. 2021;18: e1003583. doi: 10.1371/journal.pmed.1003583.

